# Provision of key components of palliative care in everyday practice in Dutch hospitals

**DOI:** 10.1101/2025.08.01.25332792

**Authors:** N. van Velzen, C.S. Heipon, M.S.A. Boddaert, H. Kazimier, Y.M. van der Linden, A.K.L. Reyners, A. Stoppelenburg, M.J.D.L van der Vorst, M.F.M. Wagemans, S. Wiegersma, L. Brom, N.J.H. Raijmakers

## Abstract

**Objective:** To examine the provision of key components of palliative care (PC) in Dutch hospitals, and to identify associated hospital- and specialist palliative care team (SPCT) characteristics.

**Materials and methods:** SPCTs from all 72 Dutch hospitals were invited to an online survey about PC practices in their hospital. The survey was conducted from January to March 2024. Data from a cross-sectional national survey among Dutch hospital-SPCTs was used. The survey included questions about hospital- and SPCT characteristics, and the current provision of key components of PC in hospitals, namely advance care planning (ACP) and routine symptom monitoring. Hospitals with and without implemented ACP and routine symptom monitoring were compared.

**Results:** In total 58 hospitals participated (81%), with a median annual admission of 20,456 inpatients (IQR 12,787) and a median SPCT referral rate of 1.4% (IQR 1.4%) of annual admissions. Routine ACP discussions were held in 58% of hospitals, with 12% consistently implementing ACP across the hospital. No differences in hospital- and SPCT characteristics were found between hospitals with or without routine ACP discussions. In 59% of hospitals, symptoms of outpatients were screened using a tool. These hospitals had both a higher SPCT referral rate (1.7% vs. 1.1%, p<0.05) and more inpatient SPCT referrals compared to those without monitoring (median of 330 (IQR 199) vs. 175 (IQR 279), p<0.05). Moreover, hospitals with routine symptom monitoring were more likely to offer an internship with SPCTs (70% vs 35%) and PC education (61% vs 17%) compared to those without (p<0.05).

**Conclusions:** Routine ACP discussions and symptom monitoring in outpatients do not yet seem common practice in Dutch hospitals. SPCTs can play an important role to further implement these key elements, together with guidelines, educational programs and a digital infrastructure for reporting and sharing outcomes.

## Introduction

As the global population continues to age and the prevalence of chronic illness increases, the need for palliative care (PC) rises (1). PC offers significant benefits for patients, including improved quality of life, reduced depressive feelings and symptom burden, and increased satisfaction with care (2–8). Additionally, PC benefits relatives by reducing stress and depression, and enhancing their satisfaction with care (9, 10).

To realise these benefits for patients and their relatives, various medical organisations have issued guidelines and recommendations for the integration of PC into standard care, including for patients with cancer or chronic obstructive pulmonary disease (11, 12). The Lancet Oncology Commission has highlighted several models to facilitate this integration, including the role of hospital-based specialist palliative care teams (SPCTs) as a key component (13). The Dutch healthcare system aims to integrate PC through a generalist-specialist model. In this model, all clinicians are expected to provide generalist PC based on their medical training, including advance care planning, basic symptom management, and supportive and end-of-life care in line with the patient’s preferences (14). PC specialists have additional expertise and broad experience in PC and can support PC generalists when needed, e.g. in case of complex symptom burden. Over the years, hospital-wide integration of the SPCTs in Dutch hospitals have progressed significantly (15). A previous study identified characteristics related to high involvement of SPCTs, such dedicated outpatient clinics and the provision of education (16). A national Dutch Delphi study on oncology hospital care identified structural implementation of advance care planning (ACP), routine symptom monitoring and the involvement of the SPCTs as key components of timely integration of PC into standard care in a generalist-specialist model (17).

For an effective and timely integration of PC into hospital care, it is essential to better understand the current provision of these key components, and how they are associated with hospital- and SPCT-characteristics. However, little is known about the current provision of ACP and routine symptom monitoring and which hospital- and SPCT characteristics are related to these practices. Therefore, the aim of this study was to examine the current provision of key components of PC in everyday practice of Dutch hospitals, and to identify associated hospital- and SPCT characteristics.

## Materials and methods

### Study design

Data from a national cross-sectional survey was used for analysis. The survey was conducted from January to March 2024 as part of a three yearly assessment of PC in Dutch hospitals. The results of the primary analysis have been reported elsewhere (18). To ensure the quality of reporting, the STROBE reporting guidelines for cross-sectional studies were adhered to (19).

### Setting and participants

Key members of the SPCTs from all 72 hospitals in the Netherlands were invited to participate in an online survey. These hospitals included general, teaching and academic hospitals, as well as dedicated cancer centres. As the respondents were no research subjects themselves and solely provided organisational data, respondent participation in the survey was regarded as implicit consent.

### Questionnaire

The questionnaire was developed as part of a three-yearly survey. The 2015 questionnaire was pilot tested for face validity, reliability, and questionnaire length by members of a SPCT (20). Each subsequent questionnaire was reviewed and updated to reflect relevant developments in the SPCTs at the time of the survey. The questionnaire included questions about hospital- and SPCT characteristics, as well as the current provision of key components of PC in hospitals. The 2024 survey, used for this study, was administered online using Survey Monkey, with email and telephone reminders sent to non-responders. Incomplete responses were followed up by email to encourage completion.

### Hospital- and SPCT characteristics

The first part of the questionnaire included items on hospital characteristics, such as hospital type, number of hospital admissions and SPCT referral rate. It also included questions about SPCT characteristics, such as number of annual inpatient SPCT referrals and the possibility for an internship with the SPCT.

### Current provision of key components of PC in hospitals

The current provision of PC in hospitals was assessed based on two key components: advance care planning (ACP) and routine symptom monitoring (17). ACP was evaluated by asking whether routine ACP discussions were conducted within the hospital (“Are advance care planning discussions conducted routinely within the hospital?”). Routine symptom monitoring was assessed by asking whether symptoms in outpatients were routinely screened using a tool (“Are symptoms routinely screened in outpatient patients using a tool?”).

### Hospital- and SPCT characteristics by key components of PC in hospitals

To assess which hospital- and SPCT characteristics are related to the current provision of key components of PC, we assessed whether these characteristics differed between hospitals with and without routine ACP discussions and between hospitals with and without routine symptom monitoring in outpatients.

### Statistical analysis

Descriptive statistics were used to summarise the hospital- and SPCT characteristics and the key components of PC. Data were presented as numbers and percentages for categorical variables and as median and interquartile range (IQR) for non-normally distributed continuous variables. Missing data on the number of hospital admissions per year were supplemented by annual reports of the hospitals. Missing data were reported for variables with more than 5% missing values. The SPCT referral rate was calculated as the number of annual inpatient referrals to the SPCT as a percentage of the number of total annual hospital admissions. Hospital- and SPCT characteristics were compared by key components of PC in univariate analyses using Chi-square tests for categorical variables and Kruskal-Wallis tests for non-normally distributed continuous variables. P-values < 0.05 were considered statistically significant. Statistical analyses were performed using STATA version 17 (StataCorp LLC, Texas, USA).

## Results

All 72 hospitals in the Netherlands were invited and 58 hospitals participated in the survey, yielding a response rate of 81%.

### Hospital- and SPCT characteristics

The participating hospitals had a median annual admission of 20,456 inpatients (IQR 11,889-24,676) and a median SPCT referral rate of 1.4% (IQR 0.8-2.2%). Most hospitals were general (43%) or teaching (40%) hospitals. The median number of annual inpatient SPCT referrals was 276. More than half of the hospitals had a dedicated PC outpatient clinic (53%) (Table 1).

**Table 1.** Hospital- and SPCT characteristics.

|  | **Total**<br>**(N=58)** |
| --- | --- |
| Number of hospital admissions / year (median, IQR) | 20,456 (12,787) |
| SPCT referral rate <sup>1</sup> (median, IQR) | 1.4 (1.4) |
| No of yearly <u>inpatient</u> SPCT referrals (median, IQR) | 276 (261) |
|  | **n (%)** |
| Type of hospital |  |
| General | 25 (43) |
| Teaching | 23 (40) |
| Academic | 8 (14) |
| Specialised | 2 (3) |
| PC assignment of the hospital executive board | 34 (61) |
| Presence of SPCT | 58 (100) |
| Presence of dedicated PC outpatient clinic | 30 (53) |
| Participation of SPCT in MDTMs <sup>2</sup> of other departments | 30 (53) |
| Presence of didactic PC curriculum <sup>3</sup> | 58 (100) |
| Combined PC and oncology educational activities <sup>4</sup> | 30 (55) <sup>5</sup> |
| Possibility for internship with SPCT <sup>6</sup> | 31 (55) |
| Continuing medical education in PC for attending oncologists | 24 (43) |
| Organisation of symposia by SPCT | 42 (72) |

<sup>1</sup> Palliative care referral rate calculated as the number of annual inpatient referrals to the specialist palliative care team as a percentage of the number of total annual hospital admissions
<sup>2</sup> *MDTM* Multidisciplinary team meeting
<sup>3</sup> Education provided to nurses, interns, residents and / or fellows hospital-wide
<sup>4</sup> Educational activities for fellows/trainees and nurses
<sup>5</sup> N=55 (3 missings)
<sup>6</sup> Possibility for residents and / or fellows hospital-wide or general practitioner trainees

### Current provision of key components of PC in hospitals

Routine ACP discussions were held in 58% of hospitals, with only 12% implementing it consistently across the hospital, 39% limiting it to specific departments, and 7% focusing on specific diagnoses. In 59% of hospitals, symptoms in outpatients were routinely screened using a tool. While physical and psychological dimensions of quality of life were almost always included in the used measurement tools (100% and 95%, respectively), the social and spiritual dimensions were addressed in only 57% (Table 2).

**Table 2.** Current provision of key components of PC in 58 Dutch hospitals.

|  | **Total**<br>**(N=58)** |
| --- | --- |
|  | **n (%)** |
| Use of instruments within hospital to identify patients with potential PC needs | 50 (86) |
| Medical specialties routinely screening outpatients for potential PC needs |  |
| Oncology | 36 (62) |
| Respiratory medicine | 20 (34) |
| Geriatrics | 18 (31) |
| Cardiology | 10 (17) |
| Internal medicine | 8 (14) |
| Neurology | 6 (10) |
| **Advance care planning (ACP)** |  |
| Routine ACP discussions within hospital |  |
| Yes | 7 (12) |
| Yes, but only for specific departments <sup>1</sup> | 22 (39) |
| Yes, but only for specific diagnoses <sup>2</sup> | 4 (7) |
| No | 24 (42) |
| ACP included as a standard component of care in the care policy | 21 (36) |
| Use of digital ACP form within hospital | 20 (35) |
| Fixed moments in the disease trajectory for ACP discussions | 10 (18) |
| Involvement of SPCT in ACP discussions | 15 (26) |
| ACP included in education for generalist clinicians provided by the SPCT | 37 (67) <sup>3</sup> |
| ACP standard topic in MDTM <sup>4</sup> of the SPCT | 37 (66) |
| **Routine symptom monitoring** |  |
| Routine symptom monitoring in outpatients using a tool | 34 (59) |
| Use of measurement tools by the SPCT | 44 (76) |
| Dimensions of quality of life included in the used measurement tool |  |
| Physical | 44 (100) |
| Psychological | 42 (95) |
| Social | 25 (57) |
| Spiritual | 25 (57) |

<sup>1</sup> Primarily for departments oncology, pulmonology, and cardiology
<sup>2</sup> Primarily for diagnoses COPD and heart failure
<sup>3</sup> N=55 (3 missings)
<sup>4</sup> MDTM Multidisciplinary team meeting

### Hospital- and SPCT characteristics by key components of PC in hospitals

Hospitals with routine symptom monitoring in outpatients had a higher median SPCT referral rate (1.7%) and more annual inpatient referrals (330) compared to those without monitoring (1.1% and 175, respectively; p<0.05). Additionally, hospitals with routine symptom monitoring were more likely to offer internships with SPCTs (70%) and continuing medical education in PC for attending oncologists (61%) compared to those without routine symptom monitoring (35% and 17%, respectively; p<0.05) (Table 3). No significant differences in hospital- and SPCT characteristics were observed between hospitals with or without routine ACP discussions.

**Table 3.**
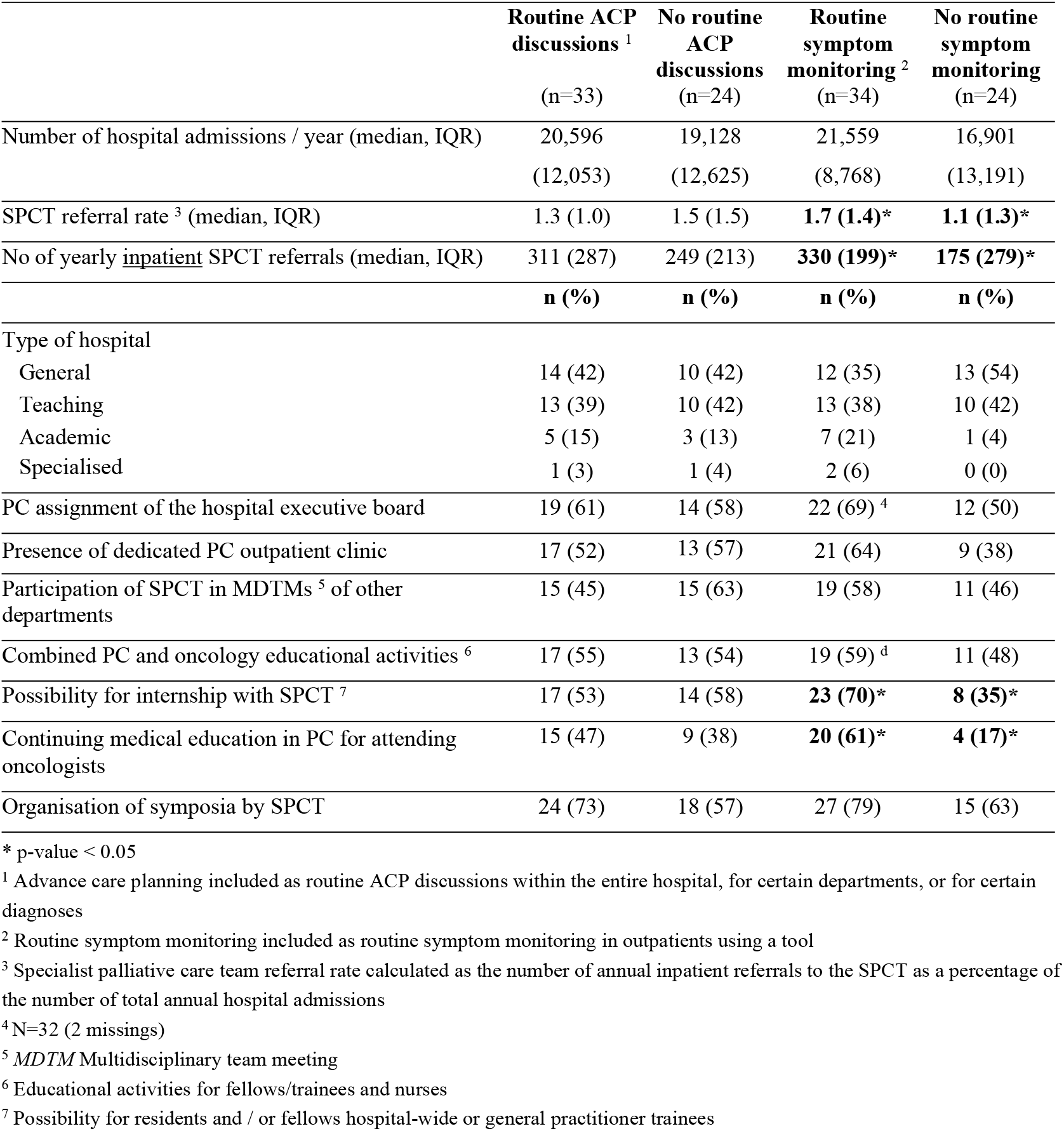
Hospital- and SPCT characteristics by key components of PC.

## Discussion

### Summary

This study examined the current provision of advance care planning (ACP) and routine symptom monitoring as key components of palliative care (PC), in Dutch hospitals. In 2023, routine symptom monitoring in outpatients and routine ACP discussions were partly implemented in hospitals. Routine symptom monitoring in outpatients was associated with a higher SPCT referral rate, offering an internship with the SPCT, and a higher likelihood to provide palliative care education for generalists. However, no significant differences in hospital- and SPCT characteristics were found between hospitals with or without routine ACP discussions.

### Advance care planning

Our findings indicate that while many hospitals are progressing toward implementing routine ACP discussions, the implementation across the entire hospital remains limited. The importance of ACP is widely acknowledged, for example by Dutch clinicians in a recent study (17), and reinforced by the National Palliative Care Program II, which identifies ACP as a core theme for the integration of PC into standard care (21). In addition, to enhance clinicians’ ability to conduct ACP discussions, the Dutch Healthcare Authority has established a dedicated payment code for Dutch hospitals, enabling clinicians to bill for ACP discussions from January 2025. ACP is associated with several benefits, such as increased use of hospice and PC services, better alignment between care preferences and the care provided, and adherence to patients’ end-of-life wishes, as well as less life-sustaining treatment and fewer hospital admissions (22, 23). The limited adoption of routine ACP discussions may reflect underlying systemic barriers. Known barriers to the implementation of ACP discussions include the absence of a standardised location for ACP documentation in the electronic health record (24). Our data show that only 35% of hospitals use a digital ACP form. Another known barrier is the lack of integration of ACP into existing clinical workflows (25). We found that only 18% of hospitals report having fixed moments in the disease trajectory for ACP discussions. Insufficient time is an additional known barrier (26). Additionally, concerns that ACP discussions may result in depression or feelings of hopelessness in patients may hinder their adoption (27). However, perceptions of involvement in ACP have been found to be positively associated with emotional functioning in patients (28).

No significant differences in hospital- and SPCT characteristics were found between hospitals with or without routine ACP discussions implemented in the workflows for specific diagnoses or departments caring for patients with life limiting diseases. We expected an association, as SPCTs may contribute to the integration of PC across the hospital by supporting and training generalists as well as developing care pathways (29). As such, a stronger association between routine ACP discussions and SPCT characteristics was expected, but the extent to which ACP is implemented across hospitals may have diluted the overall effect. Including only the hospitals with routine ACP discussions across the entire hospital (n=7, 12%) had insufficient significant power, but suggested possible associations (Supplementary Table 1).

### Routine symptom monitoring

Our findings show that in 59% of hospitals, symptoms in outpatients were routinely screened using a tool. This practice is important, as higher symptom burden can negatively impact patients’ quality of life (30, 31). To increase the practice of routine symptom monitoring it is essential to address barriers to its integration. Known barriers are tools that are burdensome for patients, their relatives and health care providers, lack of training for the use of tools as well as financial constraints (32). Additionally, effective symptom monitoring should address all four dimensions of PC - physical, psychological, social, and spiritual - as failing to do so may miss important aspects of patients’ well-being. However, our findings indicate that while physical and psychological dimensions were nearly always included in measurement tools (100% and 95%, respectively), social and spiritual dimensions were addressed in only 57% of the measurement tools used by hospitals.

Hospitals where symptoms in outpatients were routinely screened using a tool, demonstrated a SPCT that was more involved in clinical and educational activities. These hospitals had a higher median SPCT referral rate and more yearly inpatient SPCT referrals, and were more likely to offer internships with SPCTs and continuing medical education in PC for attending oncologists compared to those without routine symptom monitoring. While these practices appear to reinforce one another, the direction of influence remains uncertain due to the cross-sectional nature of our study, leading to potential reverse causality. This means that the practice of routine symptom monitoring may lead to higher SPCT referral rates since complex symptom burden is identified more often, while at the same time SPCT involvement could enhance symptom monitoring through education and training initiatives (33). Additionally, it should be noted that the higher level of educational activities in hospitals with routine symptom monitoring, could also be attributed to the fact that this group consists of more academic hospitals (21%) compared to hospitals without routine symptom monitoring (4%).

### Strengths and limitations

This is a national study to examine the current provision of key components of palliative care (PC) in Dutch hospitals and therefore provides valuable information. The survey achieved a high response rate, suggesting that our findings are likely to be generalizable to all Dutch hospitals. However, several limitations should be considered. First, the data were self-reported by members of the hospital SPCTs, which may have introduced information bias due to gaps in knowledge, as SPCT might not have been fully aware of PC practices within all departments across the hospital. Additionally, reporting bias could have occurred if respondents provided socially desirable answers to present their hospital more favourably. To minimise this potential bias, we assured participating hospitals that their responses would remain anonymous. Furthermore, while our study provides insight into the extent to which ACP and routine symptom monitoring in outpatients has been implemented in hospitals, the questionnaire did not capture information on the quality of these practices, such as the content of ACP discussions, their perceived value for patients, the documentation by clinicians, or whether there was follow-up after routine symptom monitoring (28, 34–36).

### Practical implications

The current provision of key components of PC in hospitals suggests there is potential for improvement, as not all hospitals have fully implemented ACP and routine symptom monitoring. To address this, efforts should focus on increasing awareness of PC and providing education on conducting ACP discussions, particularly among generalist PC providers. SPCTs can play a central role in advocating for the value of PC and training generalist PC providers. Facilitating this collaboration between generalist PC providers and the SPCT can foster the integration of PC across the hospital, ensuring that PC becomes an integral part of standard care. Efforts should simultaneously be directed toward improving clinical processes of PC, such as establishing clear protocols for referral pathways and standardised locations for documentation of ACP (13).

## Conclusion

While many Dutch hospitals have integrated routine advance care planning (ACP) discussions and symptom monitoring in outpatient clinics, there remains significant room for improvement. Hospitals that systematically screen outpatients for symptoms using standardised tools tend to have more engaged SPCTs in both clinical and educational activities. However, the direction of this influence remains uncertain. SPCTs can play a key role in further embedding these practices by leading educational initiatives. Overcoming common barriers—such as time constraints, insufficient digital infrastructure for reporting and sharing outcomes, and the need for clear guidelines and training programs for generalist providers—will be crucial for successful implementation. Additionally, strategic investments in SPCTs will help ensure that palliative care is consistently delivered throughout hospitals, fostering collaboration between generalist and specialist providers and ensuring that patients and their families receive timely, high-quality care. Further research is needed to evaluate the integration of ACP and symptom monitoring in daily clinical practice and to identify effective strategies for optimizing their implementation.

## Data Availability

The datasets generated and analysed during the current study are held securely by the Netherlands Comprehensive Cancer Organisation and are not publicly available due to confidentiality but are available from the corresponding author on reasonable request

